# Considering indirect benefits is critical when evaluating SARS-CoV-2 vaccine candidates

**DOI:** 10.1101/2020.08.07.20170456

**Authors:** Molly E. Gallagher, Andrew J. Sieben, Kristin N. Nelson, Alicia N. M. Kraay, Ben Lopman, Andreas Handel, Katia Koelle

**Affiliations:** Department of Biology, Emory University, Atlanta, GA, USA; School of Medicine, Emory University, Atlanta, GA, USA; Department of Epidemiology, Rollins School of Public Health, Emory University, Atlanta, GA, USA; Department of Epidemiology and Biostatistics, College of Public Health, University of Georgia, Athens, GA, USA; Emory-UGA Center of Excellence of Influenza Research and Surveillance (CEIRS), Atlanta GA, USA

## Abstract

Significant progress has already been made in development and testing of SARS-CoV-2 vaccines, and Phase III clinical trials have begun for 6 novel vaccine candidates to date. These Phase III trials seek to demonstrate direct benefits of a vaccine on vaccine recipients. However, vaccination is also known to bring about indirect benefits to a population through the reduction of virus circulation. The indirect effects of SARS-CoV-2 vaccination can play a key role in reducing case counts and COVID-19 deaths. To illustrate this point, we show through simulation that a vaccine with strong indirect effects has the potential to reduce SARS-CoV-2 circulation and COVID-19 deaths to a greater extent than an alternative vaccine with stronger direct effects but weaker indirect effects. Protection via indirect effects may be of particular importance in the context of this virus, because elderly individuals are at an elevated risk of death but are also less likely to be directly protected by vaccination due to immune senescence. We therefore encourage ongoing data collection and model development aimed at evaluating the indirect effects of forthcoming SARS-CoV-2 vaccines.

---

SARS-CoV-2 has spread globally since its emergence in December 2019 [1], resulting in more than 17 million confirmed infections and 650,000 COVID-19 deaths to date [2]. Current public health interventions aimed at curbing the spread of SARS-CoV-2 have been limited to non-pharmaceutical interventions (NPIs), including travel-associated quarantines, contact tracing, and implementation of social distancing regulations. These measures have had various degrees of success worldwide [3, 4, 5]. While critical to slowing viral spread, some of these NPIs have resulted in widespread job loss and economic hardship [6, 7, 8], and profoundly changed the way we interact with one another locally, regionally, and internationally. Given the substantial political and economic costs associated with NPIs, long-term solutions are needed. A vaccine remains the most promising solution. Thanks to tremendous research efforts worldwide, vaccine development is well underway, with more than 30 vaccine candidates in clinical trials, including 6 novel candidates in Phase III trials as of July 31st, 2020 [9].

Clinical trials for SARS-CoV-2 vaccines evaluate both the safety and efficacy of vaccine candidates. Ethically, a vaccine cannot be licensed if it does not provide a direct protective benefit to the vaccinee [10]. Direct protective benefits of vaccines include protection from infection, reduced symptom development, and lower mortality rates. While significant attention should be given to a SARS-CoV-2 vaccine’s direct benefits, vaccination can also bring about indirect effects [11]. These indirect effects decrease the infection risk of both vaccinated and unvaccinated susceptible individuals by reducing the extent of virus circulation in a community. Virus circulation can decrease because vaccinated individuals are less susceptible to infection, or because vaccinated individuals have shorter durations of infection or lower viral loads that reduce their infectiousness.

Vaccination campaigns can significantly reduce the number of infections and deaths in subpopulations that remain unvaccinated, even when vaccination coverage is quite low [12] - a crucial consideration, given that extensive vaccine coverage will be a formidable challenge. Vaccine doses, and the public health infrastucture needed to administer them, will almost certainly be limited in supply relative to demand, and given the current politically and emotionally charged climate, vaccine refusal could pose an additional barrier [13, 14]. The vast majority of the global population remains susceptible to the virus and we are likely well below the herd immunity threshold, despite the staggering rates of infection that some regions have already experienced. Therefore the indirect effects of vaccine candidates are critically important to consider when evaluating SARS-CoV-2 vaccine candidates and formulating strategies for their roll-out.

Clinical trials to test vaccine effectiveness do not evaluate population level effects. Quantifying both the direct and indirect effects of vaccines on population level outcomes, such as disease incidence and mortality, requires evaluating data after a vaccine has been in use for some time. Therefore, mathematical models are needed to assess the potential indirect effects in advance. These models can also help us gauge which vaccines should be further considered, even if direct effects may be lower than desired. To demonstrate the utility of these types of models, consider two hypothetical vaccines. Vaccine 1 reduces the risk of clinical infection in vaccinated individuals to 30% of the original risk (a direct effect), and reduces the infectiousness of vaccinated individuals to 70% of the original value (an indirect effect). Vaccine 2 reduces the risk of clinical infection to 70% of the original and the infectiousness to 30% of the original. To evaluate the direct and total (direct + indirect) effects of these vaccines, we can use a compartmental susceptible-exposed-infected-recovered (SEIR) model with infected individuals experiencing either clinical or subclinical infection (Figure 1A and supplemental material). This model classifies individuals as susceptible to infection (*S*), exposed (*E*), actively infected with either a clinical (*I_c_*) or subclinical (*I_sc_*) infection; or recovered from infection (*R*). Although not explicitly included, COVID-19 mortality can be gauged by assuming that a certain fraction of clinically infected individuals succumb to infection. In our model, we assume that susceptible individuals, once vaccinated, remain susceptible to infection but are both less likely to develop a clinical infection and less likely to transmit the virus.

Simulation of this model, in the absence of vaccination, results in a maximum of nearly 190 new clinical infections daily per 100,000 individuals, and a cumulative total of almost 9,000 clinical infections per 100k (Figure 1B). If we model the effects of distributing 40,000 vaccine doses in a population of 100k, for 40% vaccination coverage, we find that both vaccines 1 and 2 lower peak rates of clinical infections and the cumulative number of clinical infections relative to the no-vaccination scenario. However, vaccine 1 (with a greater direct effect) does not reduce the number of clinical infections to the extent that vaccine 2 (with a smaller direct effect) does. Further, vaccine 1 does not act to ‘flatten the curve’ to the extent that vaccine 2 does. The effectiveness of vaccine 2 at the population level stems from the magnitude of the indirect effects it brings about. Repeating these simulations while considering only the direct effects of the vaccines results in a less pronounced reduction in clinical infections. This result is true in both cases, but particularly for vaccine 2 (Figure 1C).

**Figure 1:**
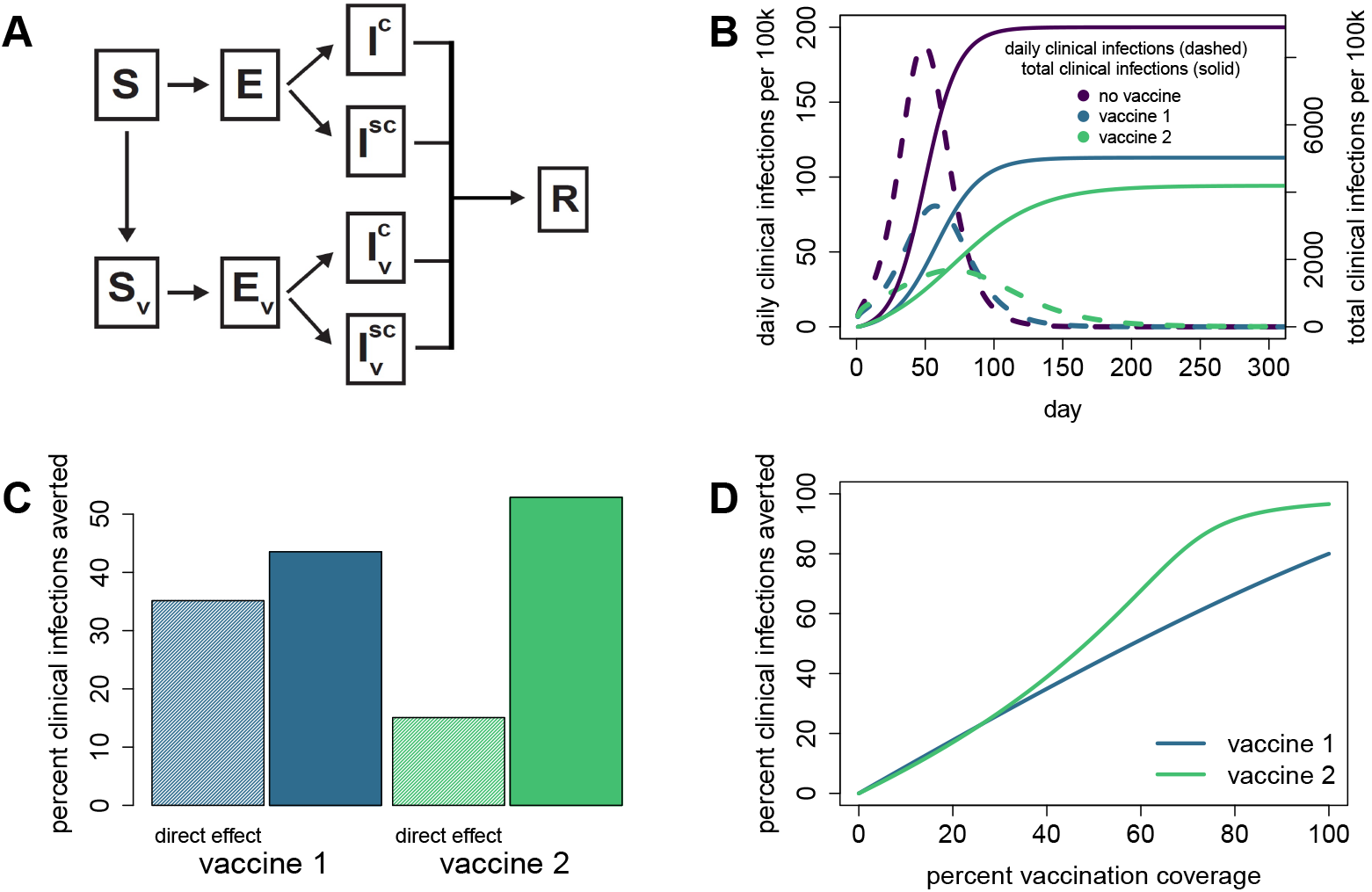
Direct and indirect effects of vaccines act together to lower population level clinical infection rates. A. Schematic of an SEIR mathematical model for SARS-CoV-2. Susceptible individuals are denoted by *S*, exposed individuals by *E*, infected individuals by *I*, and recovered individuals by *R*. Superscripts c and sc refer to clinical and subclinical infections. Subscript *υ* denotes those that are vaccinated. Model is adapted from [15]. In simulations with vaccination, vaccination of susceptible individuals occurred on day 0 [16]. B. Daily clinical infection rates and the cumulative number of clinical infections over time under a no-vaccine scenario and under separate scenarios of vaccines 1 and 2 rolled out at 40% coverage. C. Percent of clinical infections averted by vaccine 1 (blue) and vaccine 2 (green) at 40% vaccination coverage. Shaded bars indicate the percent of infections averted by the direct effect only of each vaccine. These predictions were obtained by simulating models in which vaccination does not act to reduce the infectiousness of vaccinated individuals who become infected. Solid bars indicate the total percent of clinical infections averted by each vaccine’s reduction of both clinical infection risk and infectiousness. Note that the direct effect of vaccine 1 is much greater than vaccine 2, but the total effect of vaccine 2 averts the most infections. D. Percent clinical infections averted by each vaccine for a range of vaccination coverage levels. More infections are averted by vaccine 2 than by vaccine 1 for the majority of simulated vaccination rates.

Clinical trial evaluations would heavily favor vaccine 1 over vaccine 2 because it provides a much stronger individual (direct) benefit. However, we find that significantly reducing infectiousness via vaccine 2 is as or more effective at reducing clinical infections, and therefore mortality, at the population level. The model tells us that when evaluating vaccine candidates, we should be wary of discounting a candidate that has a moderate direct effect if it also has the potential to have a pronounced indirect effect. A vaccine with relatively weak direct effects could still potentially reduce disease incidence and mortality at the population level more effectively than another vaccine that offers stronger direct protection. In the example presented here, the result that vaccine 2 averts more clinical infections than vaccine 1 is fairly robust across a range of values for vaccination coverage (Figure 1D), although the total number of infections averted by each vaccine changes significantly across coverage levels.

Our model simulations show that the reduction in clinical SARS-CoV-2 infections due to vaccination can be substantial and that indirect effects of vaccination are critically important at the population level. These indirect benefits may be particularly crucial given the severity profile of SARS-CoV-2. Indirect protection of older individuals (>65 years) may be especially important, because vaccinating older individuals is often less effective in providing direct protection due to immune senescence [17]. But quantifying the indirect effects of a vaccine is more difficult than quantifying the direct effects, and requires information not yet available. The duration of immunity, the risk of antibody-dependent enhancement, and the potential for viral evolution and vaccine escape are all unknown. While some of these effects will require years of assessment, there are some important factors that can be evaluated more quickly. For example, measurements of the level and duration of viral shedding and symptoms, from both vaccinated and unvaccinated individuals who become infected, would greatly inform our understanding of relative infectiousness. In parallel to assessing the efficacy and safety of vaccine candidates for individual recipients, we argue that research aimed at gauging indirect effects is a similarly critical area of investigation. Continued data collection and assessment of the long-term impacts of vaccines through the use of mathematical models can provide decisive information about population level protective effects. Through these efforts we can ultimately inform public health officials and politicians how to most effectively incorporate vaccination into the global public health response to SARS-CoV-2.

## Data Availability

The code to reproduce the results is hosted in a github repository which will be made public upon acceptance of the manuscript.

## Acknowledgments

The authors thank J. Baker, E. Sajewski, and the members of the COVAMOD consortium for helpful comments and discussion. The research reported in this paper was supported by NIH/NIGMS grant 3R01GM124280-03S1, NSF grant 2032084, and NIAID Centers of Excellence for Influenza Research and Surveillance (CEIRS) grant HHSN272201400004C.

## Supplementary Material

### Model Equations

We model the dynamics of SARS-CoV-2 using a set of deterministic ordinary differential equations, with susceptible individuals *S*, exposed individuals *E*, infected individuals *I*, and recovered individuals *R*. Subscripts *c* and *sc* refer to clinical and subclinical infections. Subscript *υ* denotes those that are vaccinated. Population size N is constant.

*β* represents the transmission rate (infectiousness), 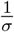 represents the average latent period, *ν* represents the proportion of exposed individuals who develop clinical symptoms, 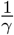 represents the average infectious period, and ρ_c_ represents the probability of death due to clinical infections.

Vaccine 1: reduces risk of clinical infection to 30% of the original value and transmission rate to 70% of the original value:

*ν_υ_* = 0.3*ν*, *β_υ_* = 0.7*β*

Vaccine 2: reduces risk of clinical infection to 70% of the original value and transmission rate to 30% of the original value:

*ν_υ_* = 0.7*ν*, *β_υ_* = 0.3*β*

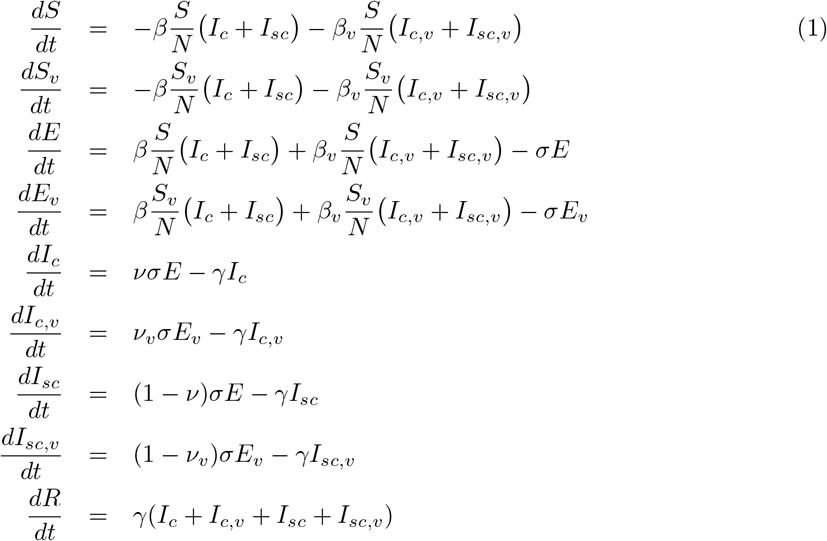

### Conditions and Parameter Values

Total population size for the simulations was fixed at *N* =100k and we assume 20% of the population is already in the ‘recovered’ class *R*. The initial size of the exposed class E was set to 200 individuals, and values for the *I_c_* and *I_sc_* classes were calculated under a fast dynamics assumption:

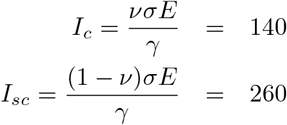

The initial size of the susceptible class *S* = 0.8(1 − *f*)*N* and the initial susceptible vaccinated class *S*_υ_ = 0.8*fN*, where *f* is the vaccination coverage level. All other vaccinated classes (*E_υ_*, *I_sc,υ_*, *I_c,υ_*) are initially set to 0, and simulations were run for one year.

We set the average latent period (1/*σ*) to 4.6 days and the average infectious period (1/*γ*) to 5 days [18]. We set the transmission rate *β* to 0.5 per day, resulting in a basic reproduction number of *R*_0_ = 2.5 [19]. We set the risk of an unvaccinated individual developing a clinical infection at *ν* = 0.14 [20], and the risk of dying from a clinical infection at *ρ_c_* = 0.02 [21, 22].

